# TESTING, TRACING AND SOCIAL DISTANCING: ASSESSING OPTIONS FOR THE CONTROL OF COVID_19

**DOI:** 10.1101/2020.04.23.20077503

**Authors:** Lia Humphrey, Edward W. Thommes, Roie Fields, Laurent Coudeville, Naseem Hakim, Ayman Chit, Monica G. Cojocaru

**Affiliations:** Department of Mathematics and Statistics, University of Guelph, Guelph, Ontario,Canada; Vaccine Epidemiology and Modeling, Sanofi Pasteur, Toronto, Ontario, Canada; Founder, covid-testing.org; Sanofi Pasteur North-America, Leslie Dan School of Pharmacy, University of Toronto, Canada

**Keywords:** Pandemic modelling, Pandemic forecasting under policy, Testing frequency policy modelling

## Abstract

In this work we present an analysis of non-pharmaceutical interventions implemented around the world in the fight against COVID-19: Social distancing, shelter-in-place, mask wearing, etc measures to protect the susceptible, together with, in various degrees, testing & contact-tracing to identify, isolate and treat the infected. The majority of countries have relied on the former, while ramping up their testing and tracing capabilities. We consider the examples of South Korea, Italy, Canada and the United States. By fitting a disease transmission model to daily case report data, we show that in each of the four countries their combination of social-distancing and testing/tracing to date have had a significant impact on the evolution of their pandemic curves. In this work we estimate the average isolation rates of infected individuals needing to occur in each country as a result of large-scale testing and contact tracing as a mean of lifting social distancing measures, without a resurgence of COVID-19. We find that an average isolation rate of an infected individual every 4.5 days (South Korea), 5.7 days (Canada) and to 6 days (Italy) would be sufficient. We also find that a rate of under 3.5 days will help in the United States, although it would not completely mitigate the second wave the country is currently under.

## 1 Introduction

In late 2019, a novel betacoronavirus called SARS-CoV-2 emerged from a live animal marketplace in Wuhan, Hubei Province, China, and has since inflicted a worldwide pandemic of a disease now referred to as COVID-19. The disease is highly contagious, with an estimated *R*_0_ between 2.2 and 4.6 [1, 2] although it is important to consider that *R*_0_ is not strictly biologically determined but rather heavily influenced by host behavioural and environmental factors [3]. The incubation period has been found to be 5.1-5.2 days, while 97.5% of patients display symptoms within 11.5 days [1, 4]. The disease spreads primarily through the respiratory tract and respiratory secretions.

As of July 16 2020, there have been a total of 13.7 million confirmed cases of COVID-19 worldwide, and over 580,000 deaths (WHO COVID-19 Situation Report 89, https://www.who.int/emergencies/diseases/novel-coronavirus-2019/situation-reports). All outbreaks, apart from that of Hubei Province continue to be active, with new cases reported daily, though a number of countries have clearly passed a (first) peak. Countries have taken various degrees of social distancing measures: lockdowns, shelter-in-place, banning gatherings, sport events, closing schools, mask wearing etc. (e.g. [5–8]) in an effort to suppress the disease, or at least prevent it from overwhelming a country’s critical care capacity. While proven effective to slow the spread, these measures have had a large effect on daily lives and economies throughout the world. Countries who managed to stave off their first wave, are now in the process of implementing (in various degrees) relaxation measures, in an effort to restore, as much and as safe as possible, their socio-economic landscapes. It is thus critical to further examine and devise strategies which will allow more phasing out of social distancing measures while preventing a resurgence of outbreaks [9], [10].

In this work, we fit a disease transmission model to daily case reports in four developed countries in order to assess the effectiveness of their COVID-19 countermeasures. We chose these due to the fact that they implemented countermeasures in various degrees and combinations, with differing outcomes to date. We begin with the examples of Italy, Canada and the United States, three countries which have relied principally on social distancing via universal shelter-in-place measures, while testing and contact tracing was implemented at an increased pace only after the pandemic was more-or-less established in their populations. We compare their pandemic evolution scenarios to that of South Korea, a country which has had and still has a tight control on COVID-19 through a combination of early aggressive testing and contact tracing, paired with social-distancing measures. We show that an analogous strategy can still provide, from this moment onward, a feasible path to further relaxing social-distancing in Italy, Canada and by extension other countries with similar pandemic profiles. The case of the United States stands apart: while coordinated large scale, frequent testing and contact tracing will help decelerate the current U.S. pandemic trajectory, this must be accompanied by a tightening rather than relaxation of social distancing measures if that country’s outbreak is to be brought under control.

The structure of the paper is as follows: in Section 2 we introduce our model main ideas, notation, and assumptions. We follow in Section 3 with the presentation of our fitted infection curves for Italy, Canada, the U.S., and South Korea, wherein we infer the net effect to date of social distancing, testing & contact tracing on the decrease of the transmission rates in each of the 4 countries. We then evaluate scenarios for countries to phase out social distancing while preventing a resurgence of COVID-19 outbreaks by assuming an increase in testing and contact tracing. We are able to derive the frequency with which a tested (infected) individual and an exposed (traced) individual need to be detected and isolated in order for each country to maintain an effective reproduction number of 1 (that is to say, each country maintains a “slow burn” of their pandemic) while assuming a major relaxation of social-distancing rules. We present a thorough discussion and conclusions in Section 5. Additional mathematical background is included in Appendix A.

## 2 Materials and methods

### 2.1 The SEIRL model

The transmission of an infectious disease in a homogeneously mixed population is often described by a Susceptible-Infectious-Recovered (SIR) model [11] or its variants, most notably a Susceptible-Exposed-Infectious-Recovered (SEIR); see [12] for a recent review. A SEIR model normalized to population size *N* is described by 4 differential equations of the form:

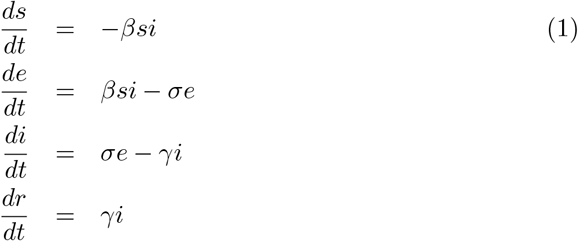

with 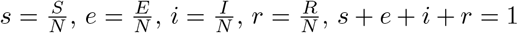 and where *β* is the rate of effective contacts, 1*/σ* = *T*_*lat*_ is the mean latent period (which may differ from the incubation period), and 1*/γ* = *T*_*inf*_ is the mean duration of infectiousness, with both times having exponential distributions. We also have the auxiliary equation for the cumulative number of cases,

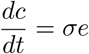

The daily incidence of cases on day *i* is then

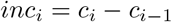

In turn, a SIR model is similar to (1) but without the *E* compartment, thus only 3 differential equations subject to *s* + *i* + *r* = 1.

The spread of an infectious disease can be halted if its effective reproduction number

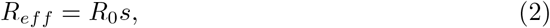

can be decreased below 1. The effective reproduction number of both the SIR and SEIR compartmental disease transmission models is

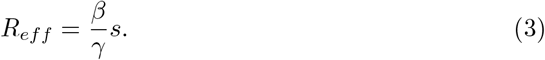

In both the SIR and SEIR model, when *s* ≈ 1 and we are near the disease-free equilibrium (1, 0, 0, 0), the early growth of both *i* and *inc* is exponential (see e.g. [12]):

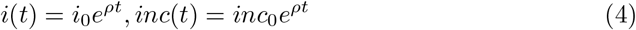

In the SIR model, the growth factor *ρ* is given by

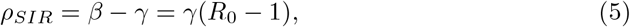

One then can express *R*_0_ in terms of *ρ*:

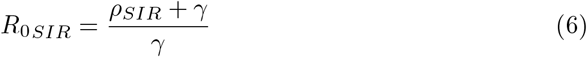

In a similar manner, in the SEIR model we have [12]:

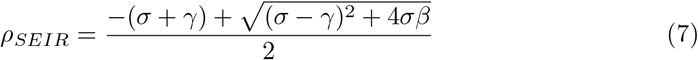

and by solving for *β* from Equation 7 we get:

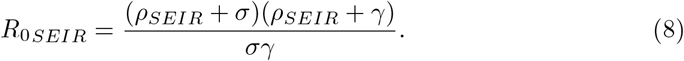

In the limit as *σ* → ∞, the SEIR model reduces to the SIR model, and accordingly, as can readily be shown by L’Hospital’s rule:

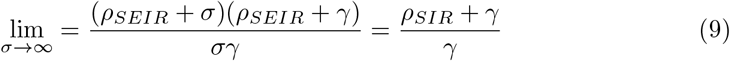

For COVID-19, as for other pandemics (e.g. SARS, MERS, the 1918 Spanish flu), we can assume the entire population to be initially susceptible. Therefore, in the early stages of an outbreak, *R*_*eff*_ ≈ *R*_0_. We will also assume that infection with COVID-19 confers subsequent immunity, which does not wane significantly over the time horizon considered here. Thus, whether they die or recover, an infected person is considered removed from the pool of susceptibles. In the absence of a vaccine or other control measures, *s*(*t*) = 1 − *c*(*t*), where *c*(*t*) is the cumulative number of people infected at time *t*.

From Equation 3, assuming *β* to be given, we see that *R*_*eff*_ can be decreased in two ways: by decreasing *s* at a rate higher than that due to infection alone; or by increasing *γ*. The former can be considered an abstraction of social distancing measures, since these effectively take a part of the population “out of circulation” as far as disease transmission is concerned. The latter can be achieved by identifying and isolating infected individuals early, thus cutting short *T*_*inf*_.

To explicitly depict the role of control measures, we adapt the SEIR model to a pandemic setting by adding an isoLated (L) compartment. As before, we include the auxiliary equation for *C*, the cumulative number of infected. The resulting SEIRL model, is described by:

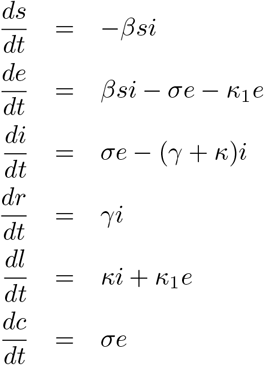

where as in the standard SEIR model, *β* is the mean rate of effective contacts, 1*/σ* = *T*_*lat*_ is the mean latent period, and 1*/γ* = *T*_*inf*_ is the mean infectious period. Finally, 1*/κ*_1_ = *T*_*isol,lat*_ and 1*/κ* = *T*_*isol,inf*_ are the mean times for the latent and infectious, respectively, to be isolated as a consequence of either testing or contact tracing.

Its effective reproduction number is then given by (see Appendix A - CHECK)

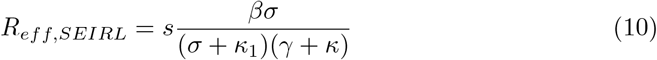

It can be shown (see Appendix A) that the exponential growth rate (Equation 4) of infected near the disease-free equilibrium is

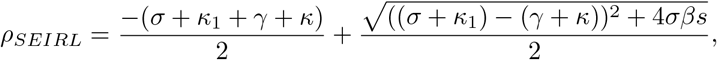

and the rate of effective contacts is

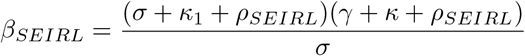

We can also express the effective reproduction number in terms of *ρ*_*SEIRL*_:

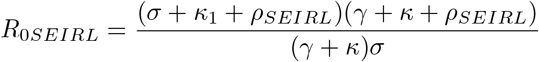

## 3 Results

### 3.1 Estimating *R*_0_ from early exponential growth

While growth is still exponential, we have from (Equation 4) that

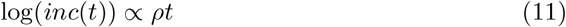

i.e. a log-linear plot of incidence versus time will have slope *ρ*. Indeed, early exponential growth can be seen to be a near-universal feature in COVID-19 daily case count data from around the world. Figures 1 and 2 plot log(*inc*) versus time for South Korea, Italy, Canada and the U.S., using time series data of daily new cases compiled by the Johns Hopkins University Center for Systems Science and Engineering (JHU CSSE) [13], retrieved from http://github.com/CSSEGISandData/COVID-19/tree/master/csse_covid_19_data

**Fig 1.**
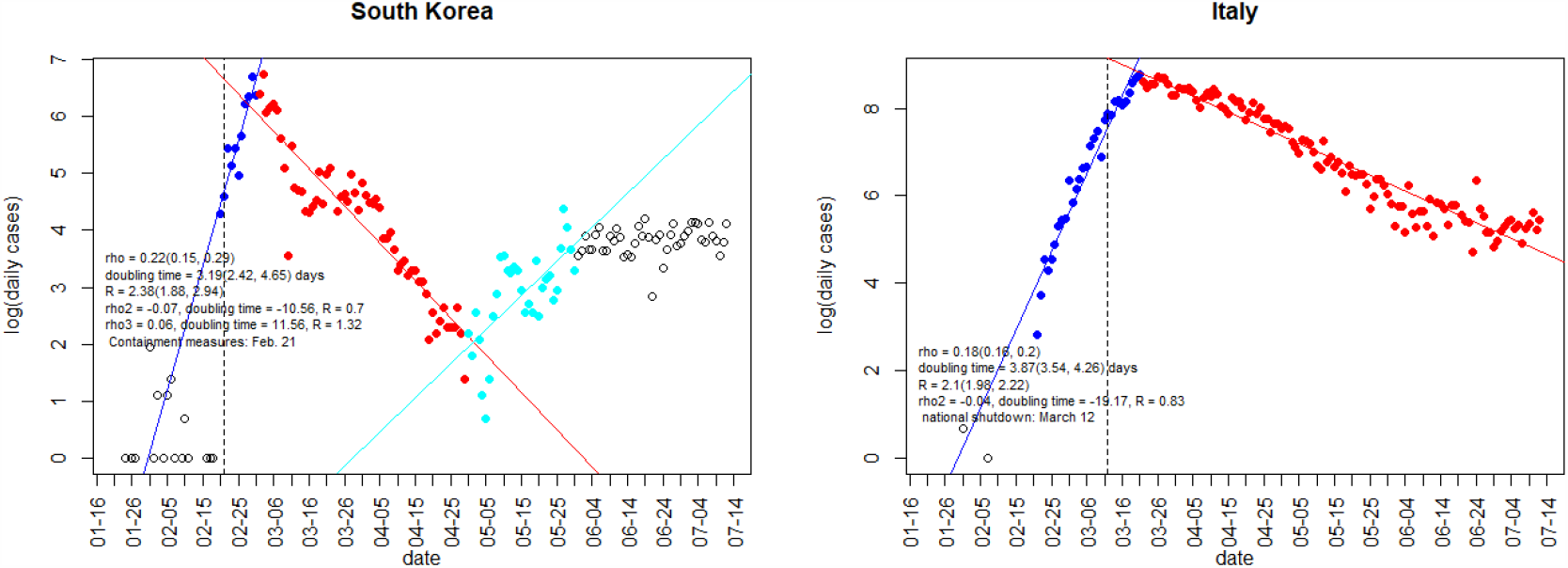
Log-linear plots of daily COVID-19 incidence versus time for South Korea and Italy. The initial linear phase corresponds to exponential growth, which subsequently turns over into sub-exponential growth. The factor *ρ* and the corresponding doubling time are estimated via a regression fit to the initial phase. *R*_0_ is calculated using Eq. 8 with *σ* = *γ* = (2.5*d*)^−1^.

**Fig 2.**
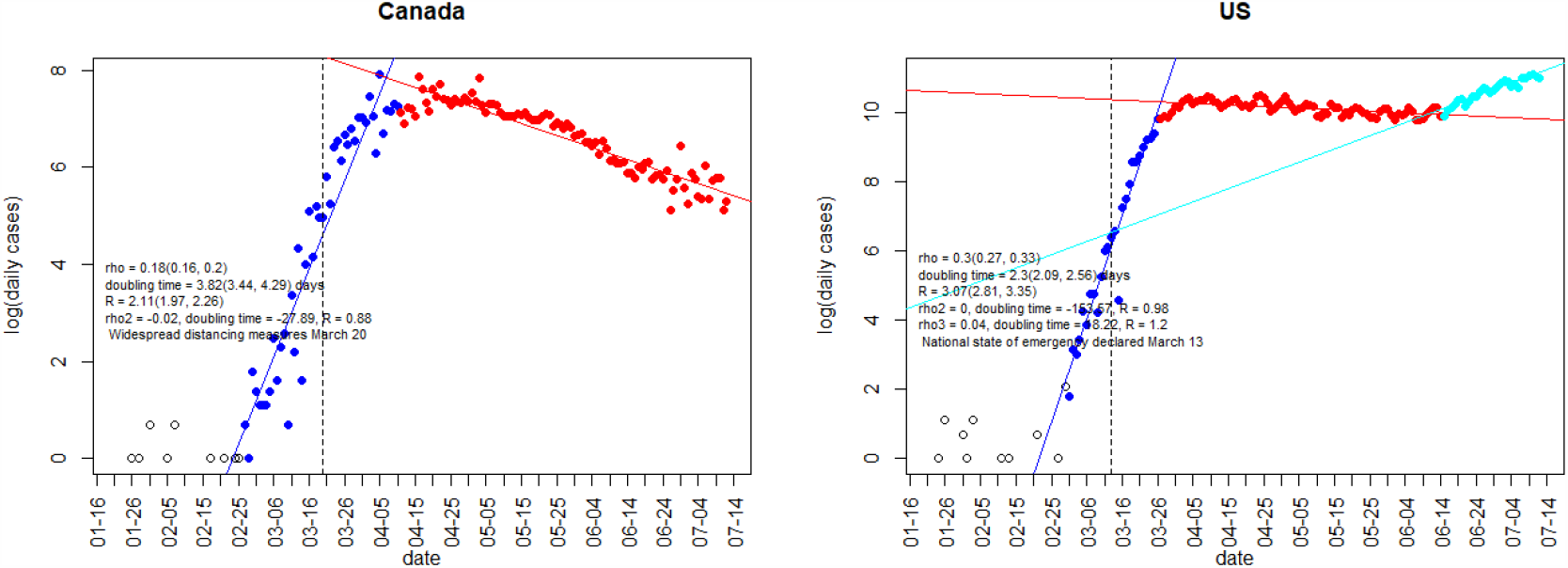
As Fig 1, but for Canada and the U.S..

In all four countries the initial linear phase is clearly apparent, followed by a transition to sub-exponential growth. This transition is sharpest for South Korea, where growth switches abruptly to decay around 1 March. Regression fit results for *ρ* and the corresponding doubling time,

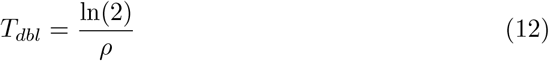

together with dates for the onset of major national-level protective measures, are given in Figures 1 and 2 and Table 1. In all four cases, the transition to sub-exponential growth occurred at or after the time that widespread protective measures were first invoked.

**Table 1.**
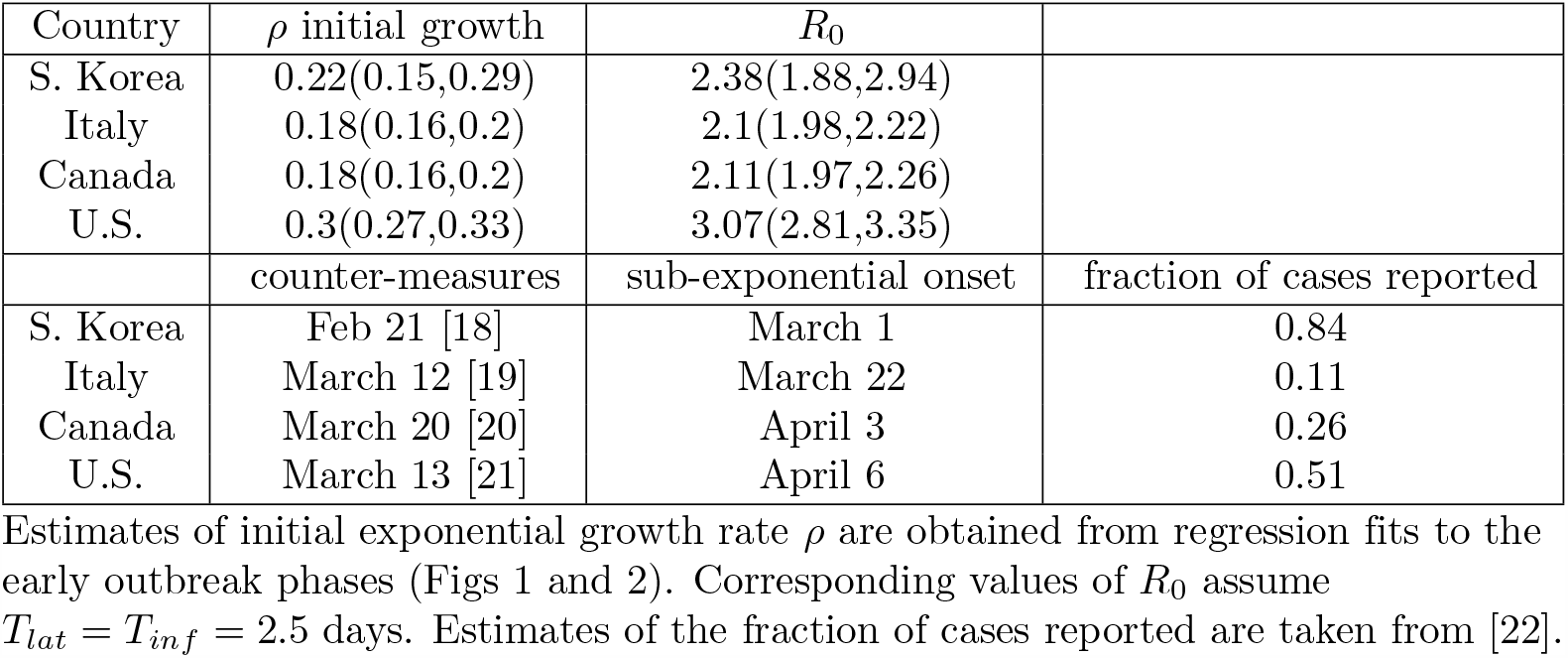
**Initial exponential growth rates and** *R*_0_ **values for South Korea, Italy, Canada, and the U**.**S**.

| Country | $\rho$ initial growth | $R_0$ | |
| --- | --- | --- | --- |
| S. Korea | 0.22(0.15,0.29) | 2.38(1.88,2.94) |  |
| Italy | 0.18(0.16,0.2) | 2.1(1.98,2.22) |  |
| Canada | 0.18(0.16,0.2) | 2.11(1.97,2.26) |  |
| U.S. | 0.3(0.27,0.33) | 3.07(2.81,3.35) |  |
|  | counter-measures | sub-exponential onset | fraction of cases reported |
| S. Korea | Feb 21 [18] | March 1 | 0.84 |
| Italy | March 12 [19] | March 22 | 0.11 |
| Canada | March 20 [20] | April 3 | 0.26 |
| U.S. | March 13 [21] | April 6 | 0.51 |

Inferring *R*_0_ from *ρ* requires choosing values for the mean latent and infectious periods. The sum of these is the mean serial interval:

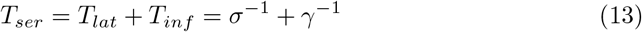

Estimates of the serial interval of COVID-19 range from 3.95 to 6.6 days [14–17]. We adopt a value of *T*_*ser*_ = 5 days. The latent period of the disease is not well constrained, but it can be shown (Appendix A) that for a given value of *T*_*ser*_ and *ρ*, the maximum value of *R*_0_ is obtained when *T*_*lat*_ = *T*_*inf*_ = *T*_*ser*_*/*2. We assume this “worst-case” scenario and let *T*_*lat*_ = *T*_*inf*_ = 2.5 days.

### 3.2 Quantifying the effectiveness of COVID-19 countermeasures thus far

We begin with the remark that all 4 countries have enacted social distancing via school closures, nationwide shutdowns, shelter-in-place orders, mask wearing, etc., all in various combinations. South Korea was the first to impose measures, followed by Italy, where the measures were ordered and coordinated eventually country-wide. In Canada most provinces enacted similar measures over the course of 1-2 weeks around March 20, 2020, while the United States has had the more heterogeneous spread of similar measures, depending on specific states.

We interpret the transition to sub-exponential growth (Figs 1 and 2) as the first signature of the effect of these measures in a given country, and use this as the starting point to infer the net effect that these measures have had up to the end of June 2020.

For each country, we fit the SEIR model solutions for daily incidence {*inc*_*model*,1_, …, *inc*_*model,n*_} to daily case reports. The model output is multiplied by a factor *kf*, where *k* is an estimate of the fraction of symptomatic cases reported, obtained using delay-adjusted case fatality rates [22], and *f* is the fraction of cases which are symptomatic, estimated to be *f* = 0.5, from a recent CDC report ^1^.

We compute − log *ℒ*, the normal negative log likelihood of the time series of observed daily incidences, {*inc*_*obs*,1_, …, *inc*_*obs,n*_}, given the model output, as a function of model parameters

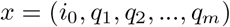

where *i*_0_ is the initial number of infected and the *q*_*i*_ are reduction factors on the rate of disease transmission, varying over time, such that 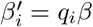 (see Table 1). *R*_0_ for each country is fixed at the respective values obtained via regression above. Parameters are drawn using unweighted (uniform) Latin hypercube sampling. The best-fit solution is the one which minimizes *L*.

We present our best fits for Italy, Canada, the U.S. and South Korea in the next figures, including:

- Linear and semi-log plots of daily incidence data of confirmed cases per country, together with maximum-likelihood model fit (“with measures, confirmed”).
- Inferred true number of infected, taking into account under-reporting and asymptomatic cases (“with measures, all”). Shown for comparison are the number of confirmed cases (“no measures, confirmed”) and all cases (“no measures, all”) expected to have occurred in the absence of countermeasures.
- Cumulative incidence and inferred reduction in effective contact rates (_*i*_*β*) due to social-distancing, mask wearing etc.
- These fits assume *κ* = *κ*_1_ = 0, in other words, the reduction in the effective contact rates 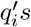 are a measure of each country’s combination of measures to date, including social-distancing, testing and tracing.

In all four countries, interventions arrested the initial exponential rise in cases and brought the effective reproduction number below. In Italy, South Korea and Canada, daily case numbers have since been brought far below their peak values.

South Korea effected the strongest and most rapid reduction in transmission. South Korea experienced a very similar early exponential growth in cases, and hence has a similar inferred *R*_0_, as the other three countries. However, its mitigation and control measures stood out from the beginning in the fact that the country employed a rapid scale-up of testing, concurrent with contact tracing and isolating of infected individuals. There are also social distancing measures imposed, but notably no shelter-in-place. Last but not least, mask wearing is a regular policy that the population adopts widely (bot only for this pandemic, but also for flu for example). Members of the population also participate in a surveillance of contacts in order to identify potential spread early. In contrast, mask wearing was instituted much later in the other 3 countries and some regions of the US are still struggling to effectively adopt it.

After some relaxation of measures in South Korea, together with a series of national holidays from April 30 to May 5 (“Golden Week”) possibly playing a role, a resurgence occurred. This appears to have since been stabilized, with (see Fig 1 above). Our fit is presented below:

## 4 Large-scale frequent testing and contact tracing as a way to relax social-distancing measures and control the spread - the SEIRL model

### 4.1 Theoretical estimates

As presented in Section 2, for a given set of values of *β, σ* and *γ*, Equation 10 gives us a closed form expression for *R*_*eff,SEIRL*_ as a function of *κ* and *κ*_1_. This relationship is depicted as a surface plot in Fig 7 for 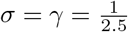 and 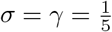. In the latter case, *R*_*eff,SEIRL*_ is nearly twice as large as in the former. It is interesting to note, though, that for both cases, the combinations of *κ* and *κ*_1_ that make *R*_*eff,SEIRL*_ = 1 (i.e. the intersections of the respective surfaces with the *R*_0_ = 1 plane) are quite similar. This can be understood as follows: As *σ* and *γ* become small compared to *κ*_1_ and *κ*, respectively, it is the latter which increasingly dominate the rate at which exposed/infected people are isolated.

**Fig 3.**
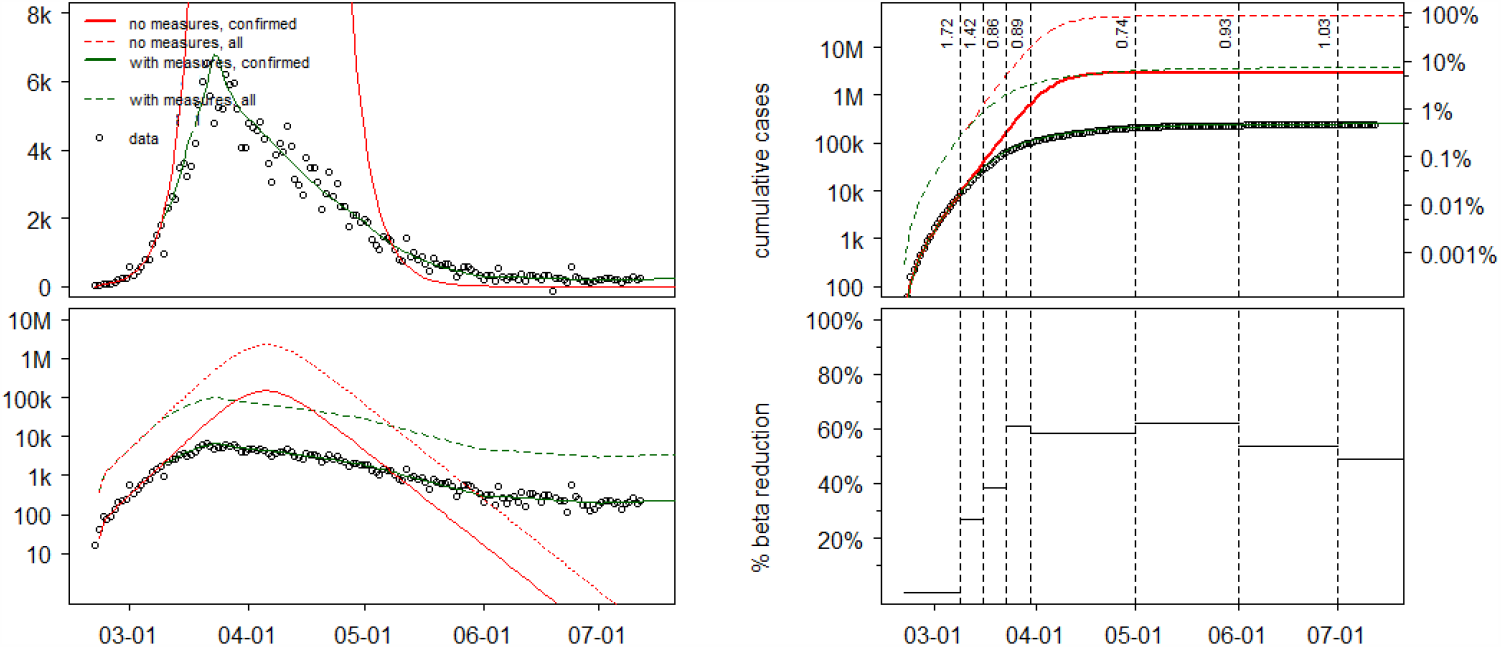
**(A) Left two panels:** Linear and semi-log plots of daily incidence data of confirmed cases in Italy, together with maximum-likelihood model fit (“with measures, confirmed”) Also shown is the inferred true number of infected, taking into account under-reporting and asymptomatic cases (“with measures, all”). Shown for comparison are the number of confirmed cases (“no measures, confirmed”) and all cases (“no measures, all”) expected to have occurred in the absence of countermeasures. **(B) Right two panels:** Cumulative incidence (top) and inferred reduction of effective contacts, together with the corresponding effective reproduction numbers (vertical numbers) (bottom).

**Fig 4.**
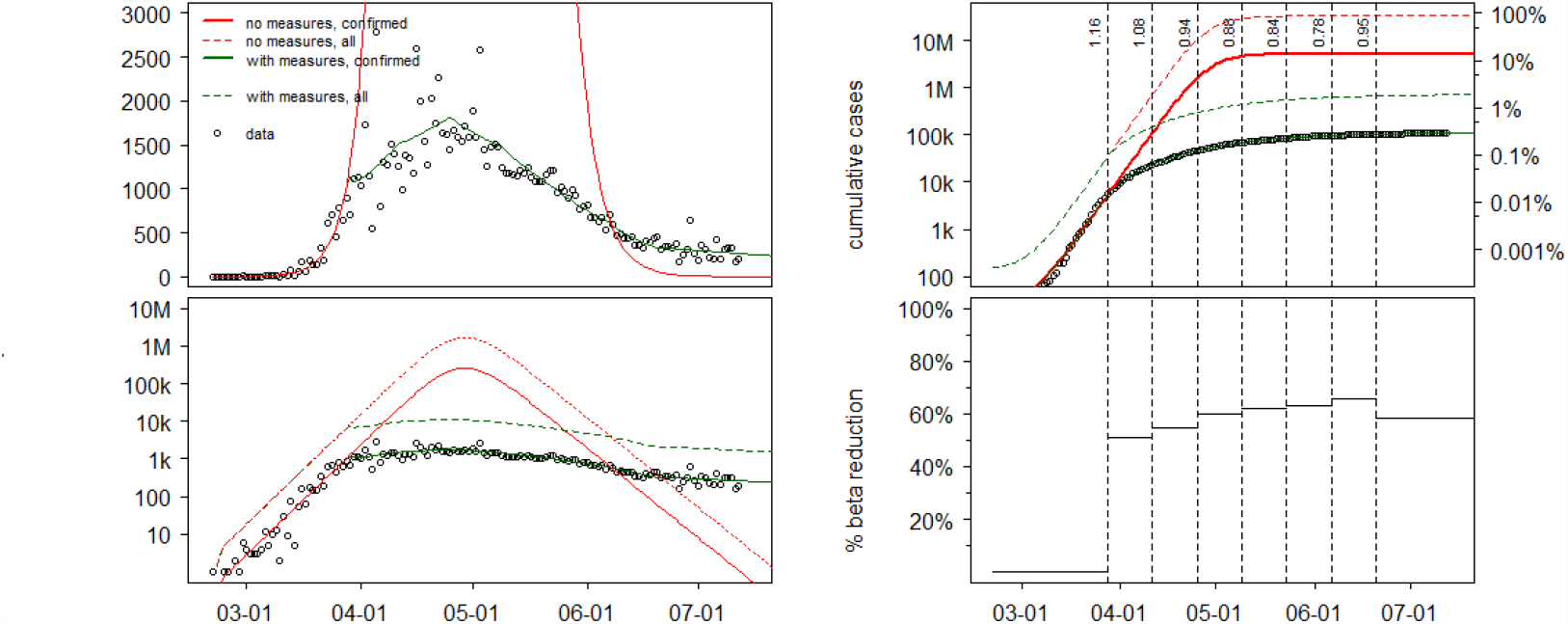
Canadian daily COVID-19 incidence (left panels). Canadian cumulative incidence fit and inferred effective contact rates reductions

**Fig 5.**
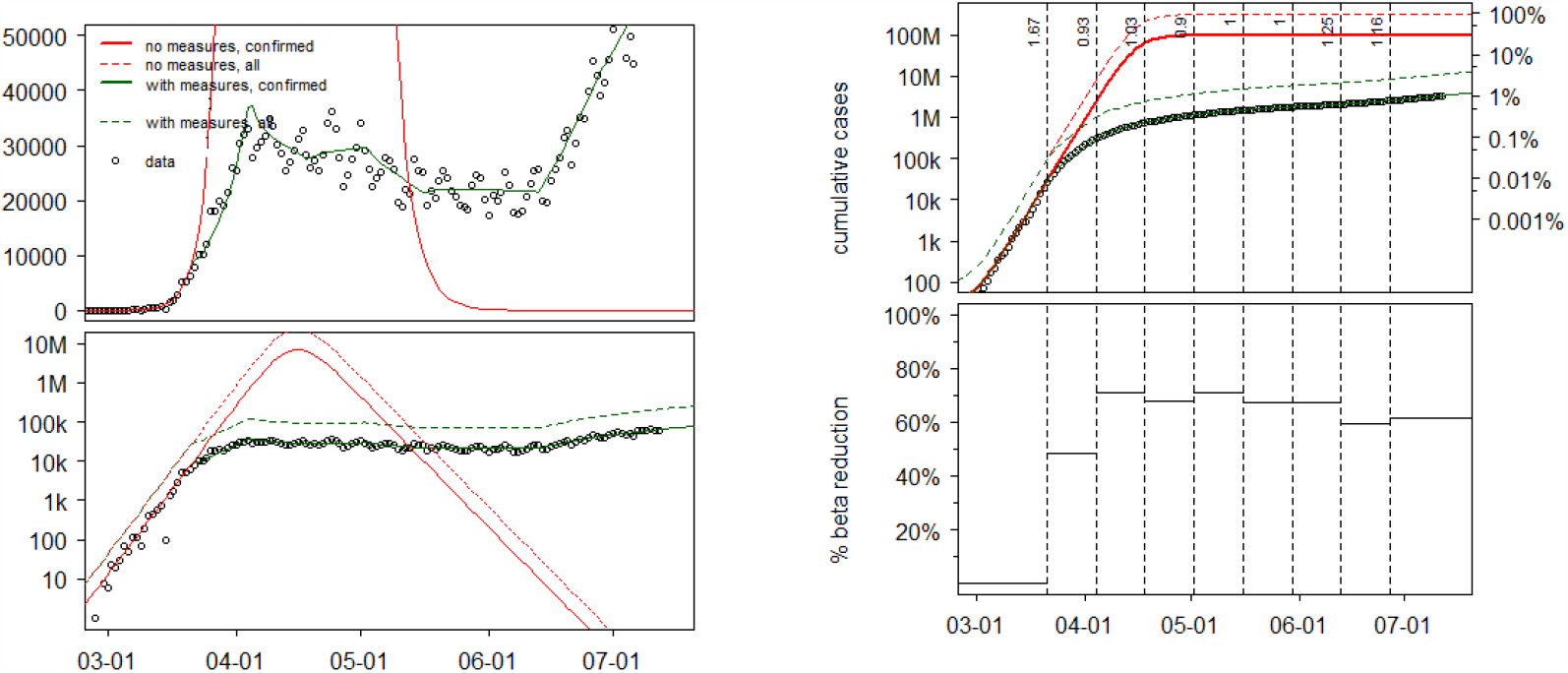
U.S. daily COVID-19 data and model fit for incidence and cumulative incidence; see caption of Fig 3 for details

**Fig 6.**
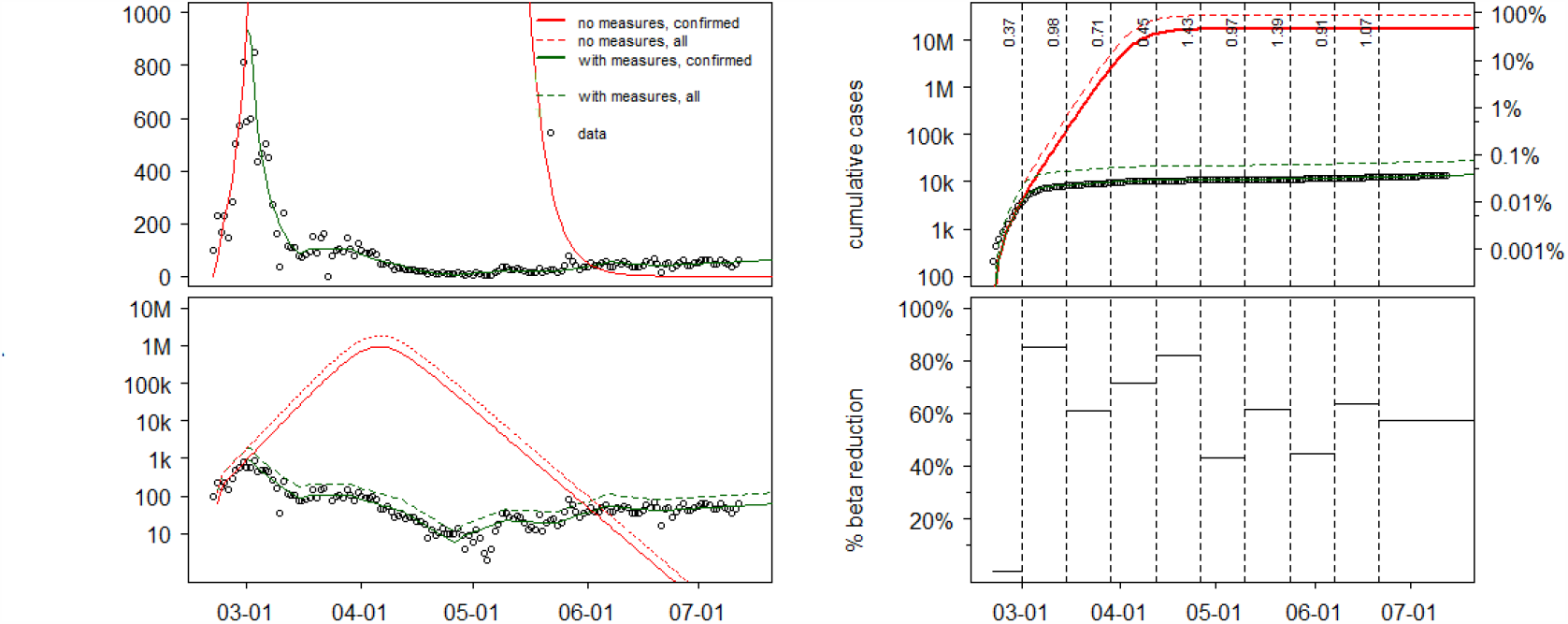
South Korea daily COVID-19 data and model fit for incidence and cumulative incidence; see caption of Fig 3 for details

**Fig 7.**
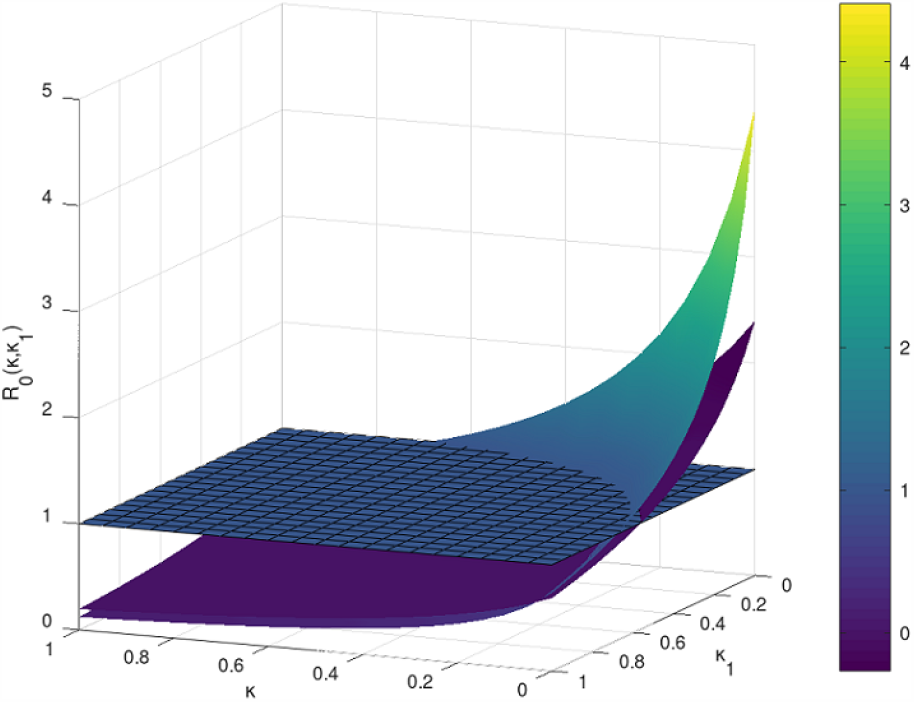
Effects of isolation rates due to testing and contact tracing on the initial value of *R*_0*S*(*Q*)*EIRL*_ model. We computed 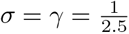 (upper most surface), 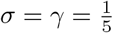 (middle surface) and the reference surface *R*_0_ = 1.

From Equation 10, we obtain the relationship between *κ* and *κ*_1_ that makes *R*_*eff, SEIRL*_ = 1:

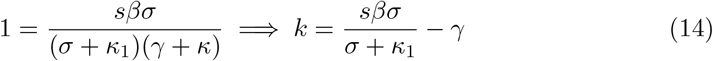

Extracting the current values of *s* from our simulations of last section, we can now compute average isolation rates due to testing and tracing that would ensure an effective reproduction number of 1, while assuming that the effective contact rates *β* will reverse to their values near the disease-free equilibrium, before any other social distancing measures were employed. We further assume that the isolation rate due to contact tracing (*κ*_1_) is the same as the isolation rate due to testing (*κ*) and we used Equation 14. We present our results compactly in Table 2 below.

**Table 2.** **Model results for South Korea, Italy, Canada, and the U.S. for *R*_*eff*_ = 1.**

| | $\beta$ | $s$ | $R_{eff}$ | $\kappa = \kappa_1$ | $(\frac{1}{\kappa}, \frac{1}{\kappa_1})$ |
| --- | --- | --- | --- | --- | --- |
| SK | 0.961 | 0.999 | 1 | 0.22 | (4.5, 4.5) days |
| Italy | 0.841 | 0.937 | 1 | 0.161 | (6.2, 6.2) days |
| Can | 0.841 | 0.982 | 1 | 0.175 | (5.7, 5.7) days |
| U.S. | 1.225 | 0.96 | 1 | 0.288 | (3.5, 3.5) days |

In all cases, we chose $\gamma = \sigma = \frac{1}{2.5}$ for a $T_{ser} = 5$ days and where $s$ are the estimated values of susceptibles remaining in each country in mid July 2020.

While the U.S. and South Korea do not seem to have an effective reproduction number under 1 at the moment, Canada and Italy have their effective reproductive number estimated to *R*_*eff*_ = 0.83, as seen from Figures 1, 2. In their cases, we can redo our estimates for the 2 countries and compute the isolation rates due to testing and tracing so that they maintain their current value of *R*_*eff*_ = 0.83 (see Table 3):

**Table 3.** **Model results for South Korea, Italy, Canada, and the U.S. for *R*_*eff*_ = 0.83.**

| | $\beta$ | $s$ | $R_{eff}$ | $\kappa = \kappa_1$ | $(\frac{1}{\kappa}, \frac{1}{\kappa_1})$ |
| --- | --- | --- | --- | --- | --- |
| Italy | 0.841 | 0.937 | 0.83 | 0.216 | (4.62, 4.62) days |
| Can | 0.841 | 0.982 | 0.83 | 0.23 | (4.33, 4.33) days |

In all cases, we chose $\gamma = \sigma = \frac{1}{2.5}$ for a $T_{ser} = 5$ days and where $s$ are the estimated values of susceptibles remaining in each country in mid July 2020. We used Eq (8).

### 4.2 Numerical results

We present next pandemic forecasts under different testing and contact tracing rates, in the four countries under consideration. We show how the theoretical estimates arise in the context of the simulated pandemic evolution in each of the 4 countries.

We depict first Canada and Italy, as they have with similar estimates, in several respective testing and contact tracing scenarios:

In Fig 9 below, We clearly see that for values of 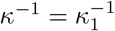 shorter than the thresholds given in Table 2, a second pandemic wave is averted in each respective country. This threshold is the least stringent in Italy, where about 6% of the population (accounting for asymptomatic and/or unreported cases) is inferred to have been infected in the first wave, as opposed to Canada.

Next we present the simulations for South Korea and the United States: We see that the isolation rates are the most stringent (lowest values in days) for the U.S., where the value of *R*_0_ inferred from the initial exponential rise of cases is right now higher than that of the other three countries (≈ 1.2). In the case of the United States, a large-scale testing and tracing operation alone will not be able to curtail the current epidemic curve around, thus strong social-distancing measures will be still be needed.

Current testing guidelines for social-distancing relaxation measures are established by the WHO in such a way that countries can relax these if positivity rates for testing are under 5% for 14 days in a row. Currently, South Korea is at 1.08%, Italy = 2.39%, Canada = 4.34% and U.S. = 6.28% ^2^ From publicly available data we have that daily testing (in numbers per day/per 1 million) are now as: Italy = 0.09823, Canada=0.085104, U.S.=0.1282 ^3^. We note that all these rates are lower than what we require in our tables above, however this is consistent with the fact that at the moment none of South Korea, Canada, Italy or U.S. have completely removed all social-distancing measures. These values give us an idea of a possible current isolation rate due to testing/tracing: 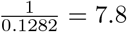 days for the U.S. and of 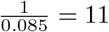 days for Canada.

## 5 Discussion

In keeping with other published findings for these and other countries, our results suggest that the COVID-19 countermeasures taken in South Korea, Italy, Canada and the United States have had a substantial impact on the course of the disease. Even accounting for estimated under-reporting, the number of cases in these countries appears thus far to have been suppressed by roughly one order of magnitude in Italy and the U.S., two orders of magnitude in Canada, and three orders of magnitude in South Korea. The development of effective vaccines and treatments is still critical to the future control of this disease, however in the interim, non-pharmaceutical interventions are the only recourse and can be effective.

Modeling studies and the United States case show that, barring a proportion of asymptomatic cases so large that the majority of people have already been infected, a second wave of disease is inevitable if distancing measures are halted or relaxed early. As shown in the case of the US, it is vital not to rush into a relaxing of distancing measures. As illustrated in Figures 9 to 10, if a change in control strategy causes *R*_*eff*_ to exceed 1, how quickly a second wave builds depends on the number of cases at the time the change has occurred. South Korea has a small number of cases, so (slightly) exceeding *R*_*eff*_ = 1 would result in a gradual climb of cases.

**Fig 8.**
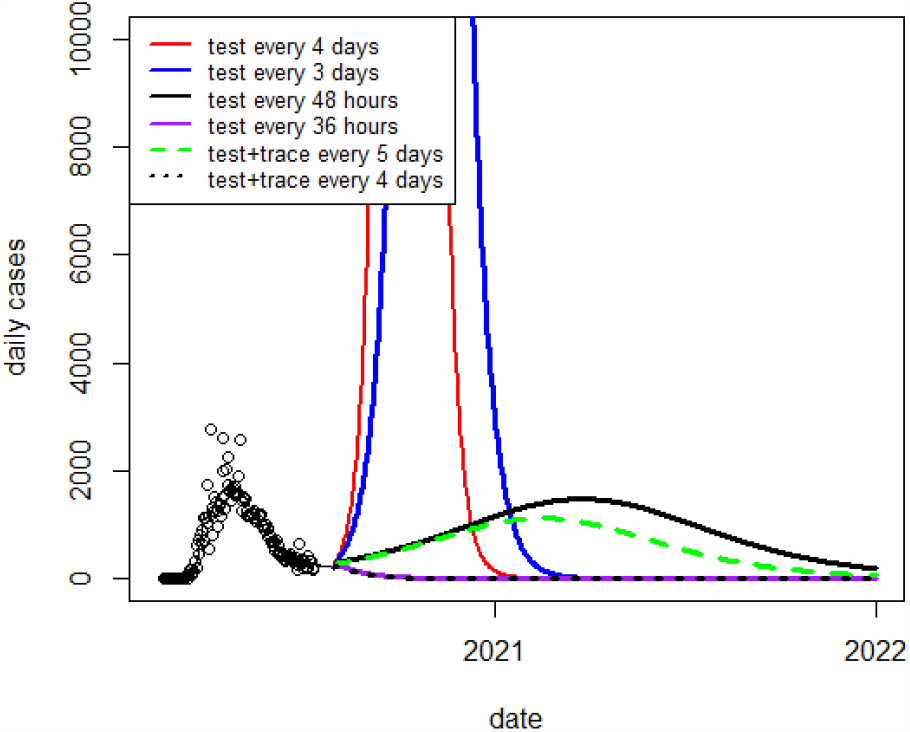
Predicted daily cases in the Canada under different rates of isolation due to testing, or testing plus contact-tracing, accompanied by a cessation of distancing measures.

**Fig 9.**
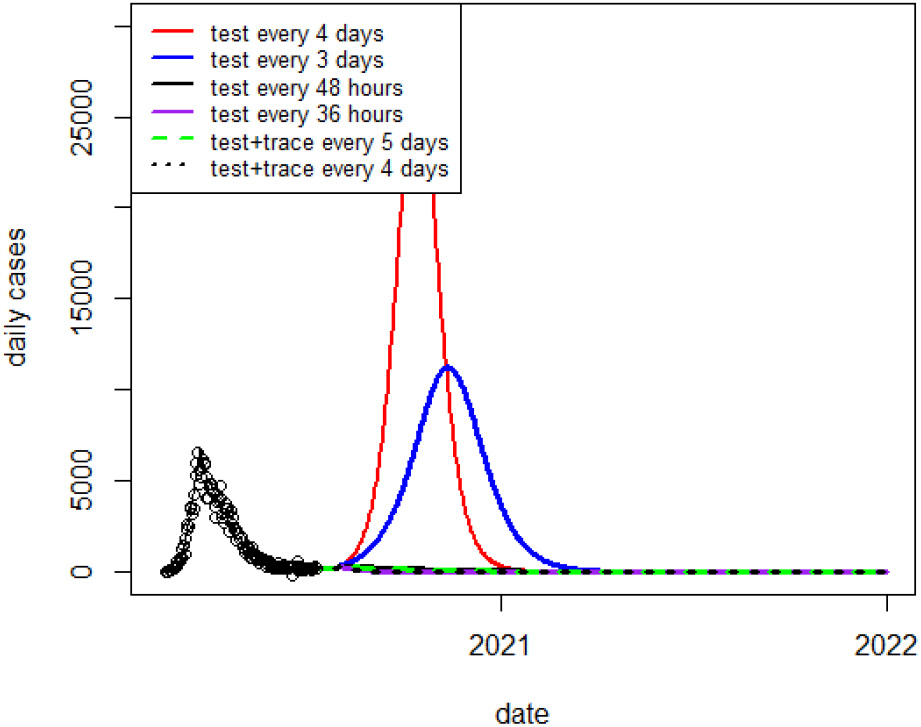
Predicted daily cases in Italy under different rates of isolation due to testing, or testing plus contact-tracing, accompanied by a cessation of distancing measures accompanied by different with various isolation rates due to testing only, or a combination of testing and contact tracing

**Fig 10.**
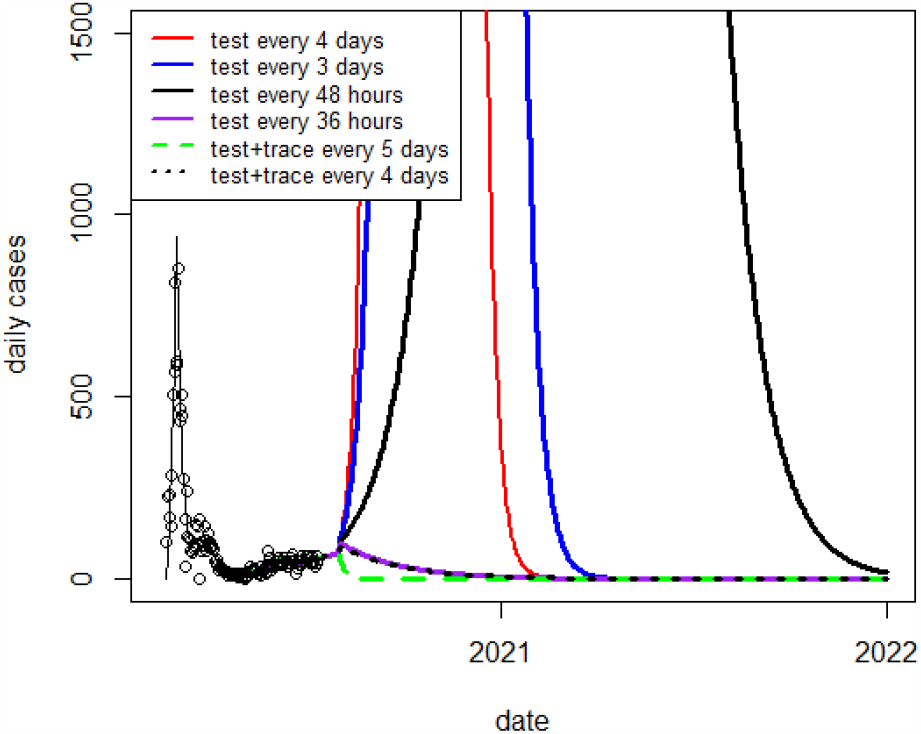
Predicted daily cases in the South Korea under different rates of isolation due to testing, or testing plus contact-tracing, accompanied by a cessation of distancing measures.

**Fig 11.**
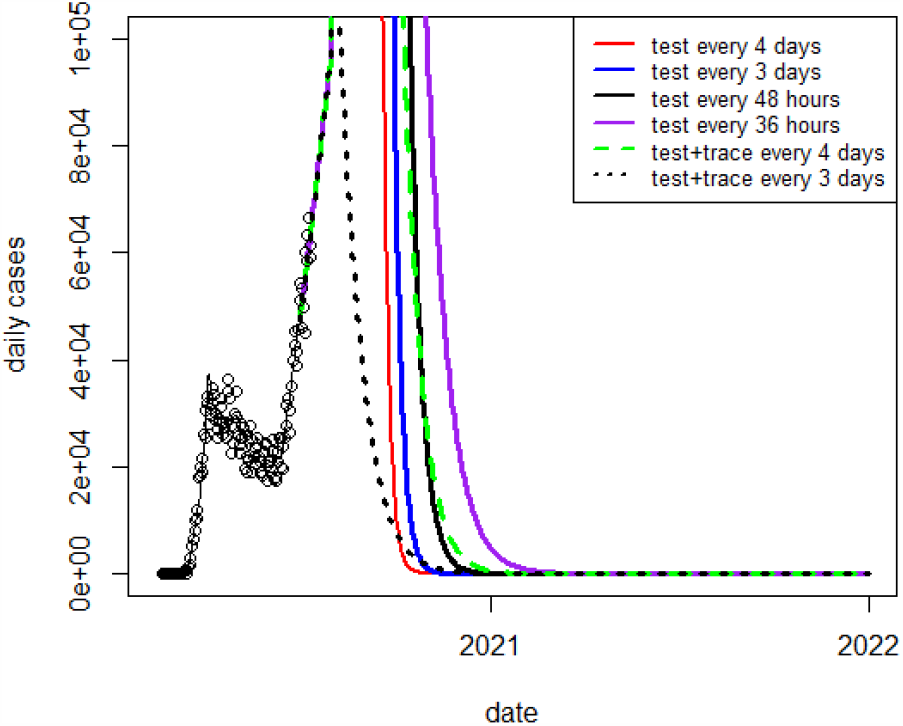
Predicted daily cases in the U.S. under different rates of isolation due to testing, or testing plus contact-tracing, accompanied by a cessation of distancing measures.

In this work, we have attempted to quantify the level of testing which would be needed to allow a country to make a near-complete return to a normal functioning of its society. Among the countries considered here, we estimate that a frequency of isolating individuals based on testing combined with contact-tracing raging from once every 6.2 days (Italy) to once every 3.5 days (U.S.) would work to keep the pandemic under a “slow-burn” control. These estimates assume a test with sensitivity at or near 100% and immediate isolation once a subject tests positive. Though reaching these targets would necessitate an undeniably large logistic effort, home-test kits availability^4^ combined with further advances in mobile device-based contact tracing ^5^ can make these strategies possible.

Our work is subject to a number of limitations. We are simulating large, heterogeneous, geographically widely-distributed populations with an unstratified disease transmission model. Although we have taken the proportion of asymptomatic COVID-19 cases to be 50%, informed by a recent CDC report, assuming asymptomatic infection confers immunity, this would mean a smaller remaining pool of susceptibles and thus a lower current effective reproduction number. Estimates of *R*_0_ from time series data of cases depend, as always, on the assumed latent and infectious periods. As we have demonstrated through (Fig 7), if these periods are longer than the isolation time, then it is the latter which principally drives the disease dynamics. Thus, our findings about threshold isolation times are relatively robust against the possibility of a substantially longer COVID-19 serial interval.

## 6 Conclusion

Testing and tracing policy directions must be strongly dependant on public cooperation and compliance. Populations will become anxious to resume more normal work, school and social schedules, while compliance with measures will become harder to enforce.

The population must be relied on to comply with self-isolation if testing positive for the virus, as well as self-isolation upon being exposed to an infected person. Last, sustainable supply chains, accuracy and reliability of possible tests as well as privacy issues around electronic contact tracing technology all present important, though not insurmountable, hurdles countries must solve. Absent universal availability of effective vaccines and treatments, the testing and tracing policies together with NPI measures are much more desirable and should be the one to strive for in the immediate short-term.

## Data Availability

All data is from publicly available sources.

## A Appendix

### A.1 Local exponential growth around a disease free equilibrium with *s*(0) ≤ 1 in an SEIRL model

We want to find a relation between the exponential growth of the infected compartment in an SEIRL model (10) and the reproductive number *R*_0_ around a disease-free equilibrium of the type 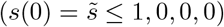) which arises as a possibility in a first wave 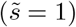 or a second wave of a pandemic such as COVID-19 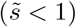.

In this case, we conduct a similar computation as in [12], but considering the 4 dimensional system of equations for *s, e, i, l* leads us to the Jacobian of the SEIRL:

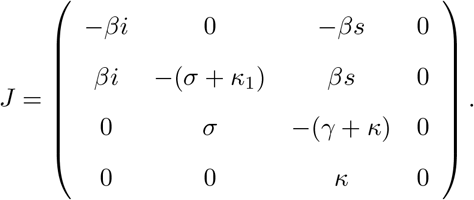

If computed at the disease free equilibrium 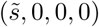 we further obtain:

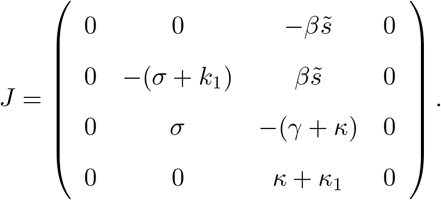

Again we note that the linearized equations for *s* and *l* are decoupled from the equations of *e* and *i*, thus, to get information on the growth rate of the infected compartment, let us try to solve the linearized reduced system in (*e, i*) based on the reduced Jacobian:

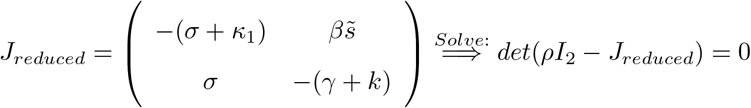

Its characteristic equation is:

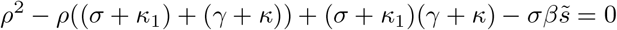

The eigenvalues of this matrix can be computed to be

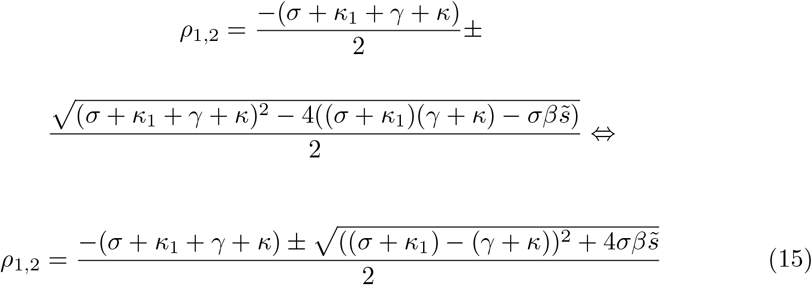

We first note that 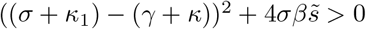, given all parameters are positive. This implies that *ρ*_1_ ≠ *ρ*_2_ *∈* ℝ and clearly *ρ*_2_ < 0. We check whether *ρ*_1_ > 0 by looking at

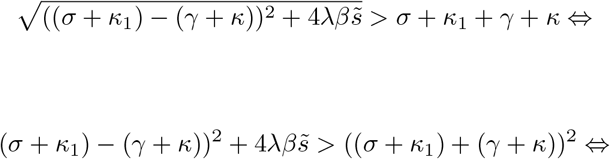

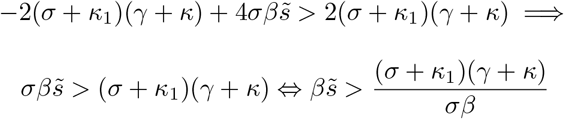

Therefore, as before, we have that

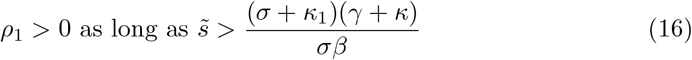

Inequality (16) simply shows that in order to not have an exponential growth from our disease free equilibrium (in other words the infection dies out), we need to allow that the initial fraction of susceptibles is lower than

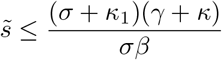

We note that *β* and *γ* are disease-dependent values on which we cannot exert control. However, *κ* and *κ*_1_ are parameters on which we can exert an exogenous control (specifically to increase them, thus raising the upper bound on fractions 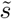 with no exponential growth in infected) which will be outlined in detail in the next section.

Continuing as in [12], we express *β* as a function of 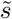, *σ, γ, κ, κ*_1_ from (15) and we get:

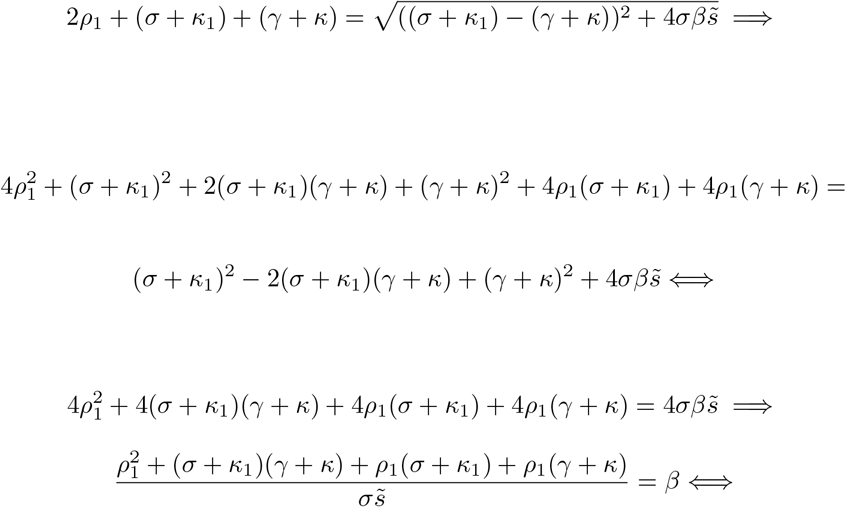

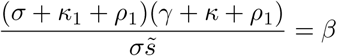

Following [23], we can use the next generation matrix to deduce *R*_0_ as the dominant eigenvalue of the next generation matrix:

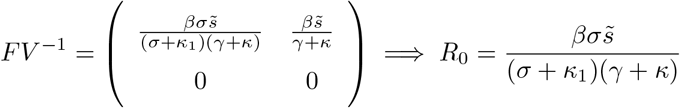

Now using the expression of *β* in that of *R*_0_ we are able to express the effective reproductive number as a function of the exponential growth and of *γ, σ, κ, κ*_1_:

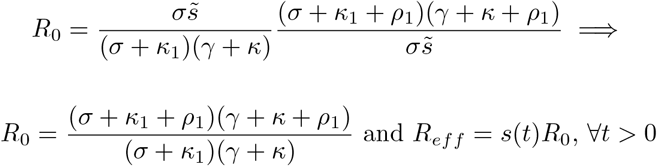

Similar to (8) we denote by *R*_0*S*(*Q*)*EIRL*_:

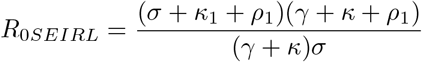

Clearly, if *κ* = *κ*_1_ = 0 then *R*_0*SEIRL*_ reduces to *R*_0*SEIR*_ in (8) of Section 2.

Let us now note that we have shown that the exponential growth factor (15), as well as the *R*_0*SEIRL*_, are dependent on the rates *κ* and *κ*_1_, that is to say, we denote by

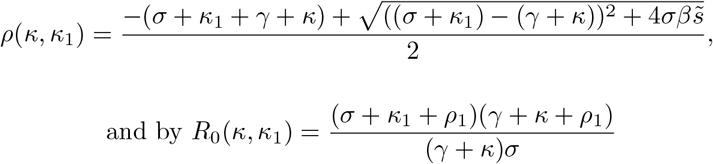

### A.2 The reproductive number as a function of *T*_*lat*_

Let us express the reproductive number, in general, as a function of 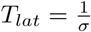 and 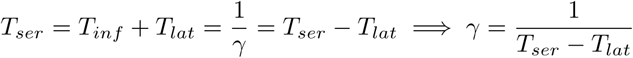

From 8 we have that

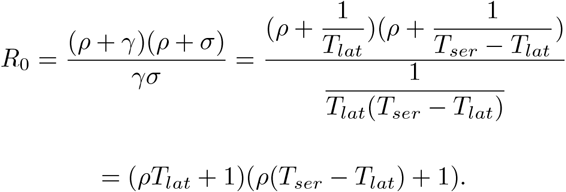

Then

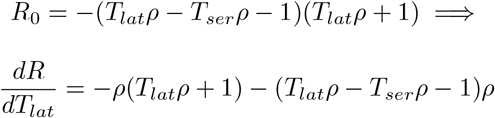

where we can solve for a *T*_*lat*_ value which maximizes *R*_0_, namely

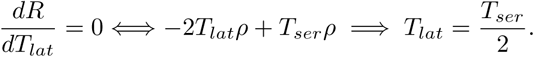

## Footnotes

1 COVID-19 Pandemic Planning Scenarios, https://www.cdc.gov/coronavirus/2019-ncov/hcp/planning-scenarios-h.pdf

2 https://coronavirus.jhu.edu/testing/international-comparison

3 https://www.statista.com/statistics/1104645/covid19-testing-rate-select-countries-world

4 A recent discussion on home-test kits this can be found in “COVID-19: A cheap, simple way to control the coronavirus” analysis https://www.nytimes.com/2020/07/03/opinion/coronavirus-tests.html

5 Canada started to test its electronic platform for volunteered participation in contact tracing in July 2020 with Ontario being first province to test it on a larger scale: https://www.cp24.com/news/covid-19-alert-app-starts-beta-testing-after-three-week-delay-1.5036434

## Notes

### Competing Interest Statement

Edward W. Thommes and Ayman Chit are employees of Sanofi Pasteur. Monica G. Cojocaru has received national research grants (NSERC CANADA) in the past, in which Sanofi Pasteur was a matching partner, however none of the work here was supported by those grants.

### Funding Statement

No third party payments have been received for this work. M. G. Cojocaru, L. Humphrey and R. Fields acknowledge support from the National Science and Engineering Research Council of Canada, Discovery Grant 400684 (PI: Cojocaru).

